# A cross-sectional study of COVID-19 knowledge, beliefs and prevention behaviors among adults in Senegal

**DOI:** 10.1101/2021.09.29.21264312

**Authors:** Matthew D. Kearney, Marta Bornstein, Marieme Fall, Roch Nianogo, Deborah Glik, Philip M. Massey

## Abstract

**Background:** COVID-19 is an ongoing threat to global public health since its emergence at the end of 2019, yet little is known about how populations in francophone West Africa have responded to COVID-19 in their daily lives. Senegal, in particular, has been noted for its relative success in mitigating the spread and impact of COVID-19. We report original research findings on COVID-19 beliefs and prevention behaviors in a sample of Senegalese adults.

**Methods:** A multi-modal cross-sectional study was conducted to describe COVID-19 beliefs and prevention behaviors in a sample of Senegalese adults and to identify potential predictors of prevention behaviors. Univariate, bivariate, and multivariate statistics were generated to describe the sample and explore potential correlations.

**Results:** Mask wearing, hand washing, and use of hand sanitizer were most frequently reported. Social distancing and staying at home were also reported albeit to a lower degree. We also identified a range of psychosocial and demographic predictors for COVID-19 prevention behaviors. Men, compared to women, had lower odds (OR=0.59) of reporting prevention behaviors. Rural residents (vs. urban; OR=1.49) and participants with at least a high school education (vs. less than high school education; OR=1.33) were more likely to report COVID-19 prevention behaviors.

**Discussion:** Stakeholders and decision makers in Senegal and across Africa can use place-based evidence like ours to address COVID-19 risk factors and intervene effectively with policies and programming. Use of both phone and online surveys enhances representation and study generalizability and should be considered in future research with hard-to-reach populations.

**Article Summary:** *Strengths and limitations of this study:*

- The main strength of our study is the use of a multi-modal data collection strategy, online and via telephone. Had we relied on a single method, our sample’s demographic characteristics would likely have differed while also introducing selection bias.
- Our recruitment strategy may have also increased the potential for selection bias because participants were recruited online and on-the-ground in Senegal; thus, all participants, by the nature of the recruitment methods, had access to the internet and/or a cell phone. To address potential confounding between recruitment methods, we controlled for recruitment method in our multivariate regression modelling.
- Prior to our study, little evidence was available about how populations in francophone West Africa have responded to COVID-19 in their daily lives. We adapted pre-existing research panels and developed novel data collection instruments to capture information about COVID-19, and this may serve as a model for the responsiveness of ongoing scholarship to future global events.
- A study with a larger sample may have been able to identify relationships between knowledge and behaviors, and our study was not design with enough statistical power to detect significant differences.
- The findings of our study may not be generalizable beyond Senegal, or more specifically Senegalese adults.

## Introduction

COVID-19 is an ongoing threat to global public health since its emergence at the end of 2019, yet little is known about how populations in low-resource settings have responded to COVID-19 in their daily lives. Although research is emerging about COVID-19 infection, strategies for prevention, and risk factors for severe disease and mortality, there is a dearth of literature about COVID-19 beliefs, specifically in the West Africa region. For example, between January and September of 2020, just 4.3% of research articles about COVID-19 were focused on Africa or a specific African nation.[1] Given that ongoing prevention strategies implemented at the country, community, and individual level will be needed to curb the COVID-19 pandemic, it is critical to study health beliefs and adherence to prevention behaviors within specific social and political contexts.[2]

Providing relevant and timely research is essential for guiding evidence-based outreach and interventions, especially during public health emergencies. It is particularly important to advance equity within public health, and the health of populations, by understanding the social contexts and health beliefs among indigenous, marginalized, and vulnerable populations around the globe.[3,4] The current study provides timely, critical, and actionable information about COVID-19 beliefs and prevention behaviors for stakeholders, policy makers, and public health programs to reduce the burden of COVID-19 in West Africa by reporting on findings among a sample of adults from the West African nation of Senegal.

### West African Context

West Africa is a distinct language and cultural region made up of 16 sovereign nations, many of which were formerly French and British colonies. Increasing internet infrastructure and use of mobile technologies has bridged historical digital disparities and inequities by allowing unprecedented access to health information for many peoples living in West Africa and other developing regions.[5–7] Francophone West Africa, in particular, is underrepresented in global health research because of language barriers to publication and collaboration, leading to more research in English-speaking nations like Nigeria and Ghana.[8–10] Increasing global internet connectivity has also augmented representation of formerly hard-to-reach vulnerable and marginalized populations in the research process as both participants and collaborators.[8–10]

Senegal is the former capital of French West Africa and home to over 16 million people. Approximately 90% of the population is Muslim and 5% are Christian, most of whom are Catholic.[11] Senegalese culture and politics are infused with both French and Islamic influences, and Senegal is often considered a model democracy. Although Senegal has a thriving tourism industry and developed financial services and international trade sectors, the nation still ranks low in the United Nation’s Human Development Index, 35^th^ out of the 53 ranked African nations.[12]

### COVID-19 in West Africa

Compared to the burden of COVID-19 in Europe, the Americas, the Middle East and South Asia, the burden of COVID-19 in Africa has been quite low. Relative to the rest of the world, Africa defied initial expectations by experiencing the lowest burden of reported COVID-19 cases and deaths.[13,14] Questions remain about why COVID-19 infections are reportedly low in Africa, in particular in West Africa where there have been 377 cases per 100,000 persons cumulatively. Although a recent analysis by Aduh and colleagues (2020) reported on perceptions and behaviors towards COVID-19 across Sub-Saharan Africa, we have much to learn from populations in densely populated West Africa, a comparatively more urban Sub-Saharan region home to more than 400 million people.[16]

Some researchers have questioned the accuracy of African COVID-19 case numbers,[13] hypothesizing that the burden of disease could be much higher than reported. Uncertainty about the true trajectory of COVID-19 in Africa may leave all continents vulnerable to future outbreaks, particularly regions with low herd immunity due to lack of exposure and vaccine resource constraints. It is likely that places that that have not yet needed to adopt strong prevention efforts because of reportedly low case numbers therefore may be especially vulnerable to future outbreaks. Thus, despite relatively low case numbers, it is still vital to investigate COVID-19 health beliefs and prevention behaviors in West Africa peoples to identify potential correlates and predictors of viral prevention behaviors.

### Study Objectives

To explore COVID-19 beliefs and prevention behaviors in francophone West Africa, the current study analyzes survey data collected from a sample of adults in Senegal. The survey, which was administered both online and via telephone, included a module on COVID-19 behaviors and beliefs. Specifically, this study applies constructs from the Health Belief Model to answer the following questions:[17,18]

1. In Senegal, what is the level of knowledge and awareness about COVID-19?
2. To what extent is COVID-19 perceived as a health threat?
3. What is the relationship between COVID-19 beliefs and prevention behaviors?

## Methods

### Study Design, Participants and Setting

From June-August 2020, we conducted a multi-modal cross-sectional survey that included questions regarding COVID-19 beliefs and prevention behaviors among adults in Senegal. The survey was administered via the internet and telephone, which was appropriate given the constraints of research during COVID-19. Respondents were participating in a separate ongoing longitudinal cohort study.

### Data Collection

This survey used two modes of data collection: online and telephone. The telephone survey used a random selection from a panel of phone numbers of individuals who lived in Senegal and agreed to participate in media-related research. A quota system was used to match the overall population of Senegal on gender and have an even distribution of age and level of education. A total of 1,221 participants were enrolled in the study via telephone, split approximately equally between participants living insider versus outside of Dakar, Senegal and by gender. Online participants from Senegal were recruited using social media advertisements on Facebook and YouTube. All online participants were part of a cohort study about entertainment education in West Africa that began in late summer 2019; of the 1,286 cohort participants recruited online in summer 2019, 231 both were eligible to participate (i.e., Senegalese adults) and completed the COVID-19 questionnaire items. Through our multi-modal strategy, responses about COVID-19 were collected from 1,452 Senegalese adults.

For both online and telephone participants, all individuals were required to be 18 years or older. Potential participants were informed about the study and provided consent to participate in the survey (online) or interview (telephone). Consenting participants were compensated with 2000 Central African CFA franc (∼4 USD) for their time. Incomplete responses with missing data were excluded. All study procedures and protocols were reviewed by and approved or determined to be exempt by institutional ethical review boards.

### Measurement

Survey items included a series of self-reported demographic questions (e.g., age, education) as well as a 15-item section specifically about COVID-19 (see Appendix A). The COVID-19 section contained three measurement domains: 1) knowledge & awareness, 2) perceived threat to health, and 3) prevention behaviors. COVID-19 items were developed based on available guidance from the WHO and CDC and were reviewed by French, English, and Wolof speaking team members. Response options were binary (e.g., agree/disagree) or Likert-style (i.e., ordinal). Our data collection instrument drew upon the Health Belief Model (HBM),[18] a behavior change framework for public health practice positing that knowledge of a potential negative outcome and perceived threat of that outcome precede behavior change.[17] Other recent studies have similarly relied on HBM constructs to explore COVID-19.[19–21] COVID-19 response items were aggregated into distinct scales based on the three measurement domains. Each scale ranged from 0 to 15. As the COVID-19 items were novel, these “scales” were used as a proxy measure for determining the degree of knowledge and awareness, perceived risk, and practicing of known prevention behaviors including 1) handwashing, 2) using hand sanitizer, 3) wearing a mask, 4) social distancing, and 5) staying at home.

### Data Analysis

Univariate and bivariate statistics were generated to describe the sample and explore potential correlations. Chi-squared tests assessed differences in demographic attributes between the online and telephone collection methods. We used Pearson’s correlation coefficient to explore relationships between items in the COVID-19 scales. For multivariate analysis, we recoded level of education, marital status, location (e.g., urban/rural), religion, and occupation to reduce the number of categories, creating binary or ordinal variables, as shown in Table 1. Participant age was collected as a continuous measure and recoded as both quartile age groups and as binary (above versus below median age). Median values were calculated for each of the three COVID-19 scales and used as thresholds to create binary variables (e.g., 0=low knowledge;1=high knowledge).

**Table 1.** Sample characteristics. N=1,452. Data collected via phone and online surveys from Senegalese adults between June-August 2020.

| Characteristic | Category | # | % |
| --- | --- | --- | --- |
| Survey modality | Telephone | 1,221 | 84.1 |
|  | Online | 231 | 15.9 |
| Age Quartile | <21 | 353 | 24.3 |
|  | 22-25 | 352 | 24.2 |
|  | 26-34 | 354 | 24.4 |
|  | 35+ | 393 | 27.1 |
| Sex | Female | 752 | 51.8 |
|  | Male | 700 | 48.2 |
| Location | Urban | 1,118 | 77.1 |
|  | Rural | 291 | 20.1 |
|  | Urban + Rural | 42 | 2.9 |
| Education | Superior | 381 | 26.2 |
|  | Secondary | 344 | 23.7 |
|  | Primary | 284 | 19.6 |
|  | Middle | 231 | 15.9 |
|  | No | 145 | 10.0 |
|  | Other | 67 | 4.6 |
| Marital status | Single | 804 | 55.4 |
|  | Married - monogamous | 460 | 31.7 |
|  | Married - polygamist | 161 | 11.1 |
|  | Divorced | 20 | 1.4 |
|  | Widower | 7 | 0.5 |
| Occupation | Informal sector | 569 | 39.2 |
|  | Student | 400 | 27.6 |
|  | Formal sector | 225 | 15.5 |
|  | Other | 112 | 7.7 |
|  | Housewife | 96 | 6.6 |
|  | Retired, out of work | 49 | 3.4 |
|  | Religious leader | 1 | 0.1 |
| Religion | Muslim | 1,372 | 94.5 |
|  | Catholic | 65 | 4.5 |
|  | Protestant / Evangelical | 8 | 0.6 |
|  | No | 3 | 0.2 |
|  | Other | 3 | 0.2 |
|  | Traditionalist | 1 | 0.1 |

We built five logistic regression models that predicted COVID-19 behaviors individually (i.e., hand washing, mask wearing, social distancing, staying home) and in aggregate, the results of which are presented as Forest plots (Figure 1). For individual behavior models, dependent variables were whether or not participants practiced prevention behaviors daily in the past week; for the aggregate model the dependent variable was the prevention behavior scale ranging from 0 (practice no behaviors in past week) to 15 (practice all behaviors daily). Independent variables in each model included demographic items, perceived threat (high/low), and knowledge & awareness (high/low). All statistical analyses were conducted using Stata IC software (version 15). Forest plots were generated using the ‘forestplot’ package in RStudio software.[22]

**Figure 1.**
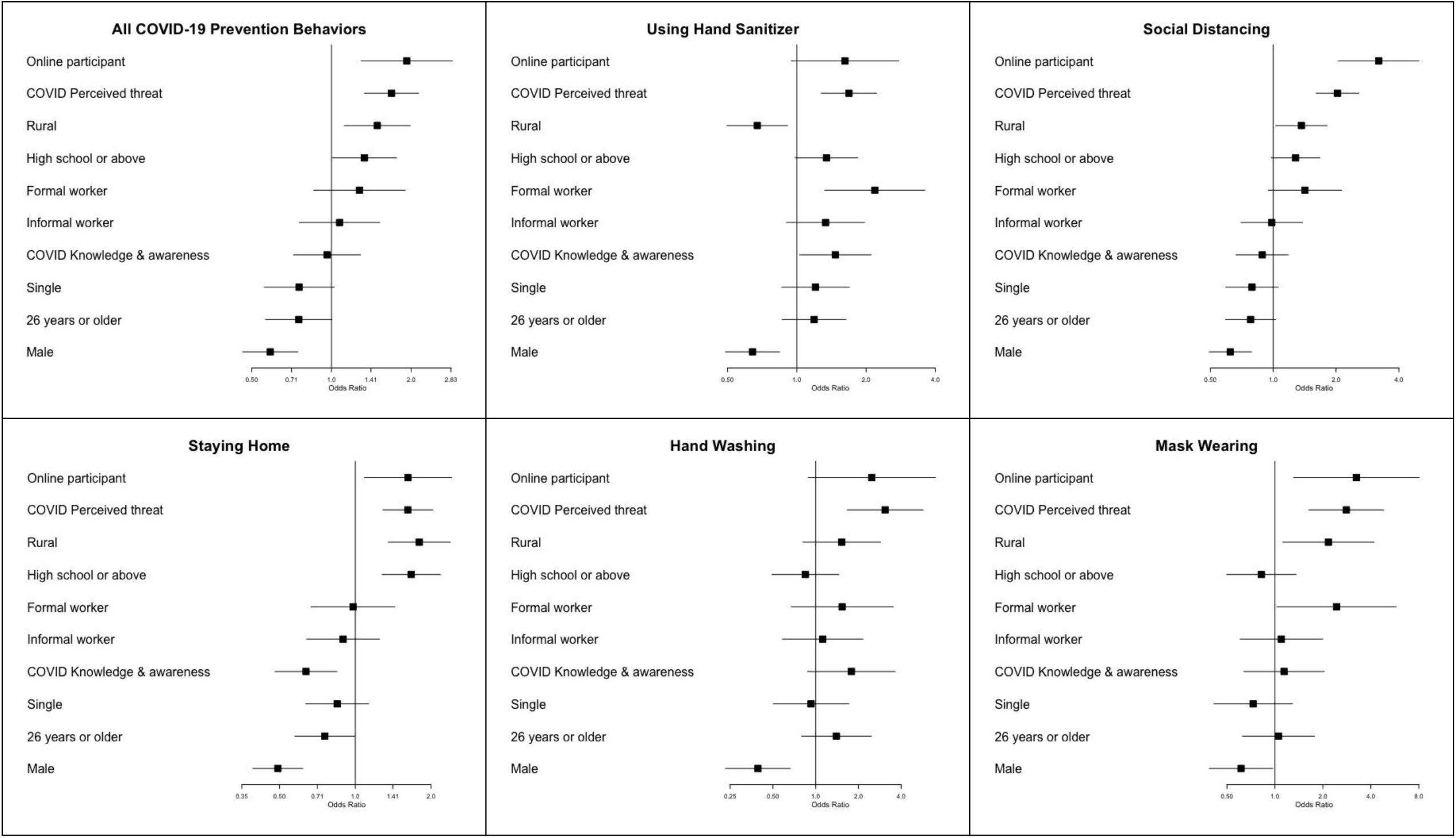
Forest plot diagrams for logistic regression models predicting COVID-19 prevention behaviors (aggregate + individually). Square box represents point estimate and horizontal bar represents 95% CI. Confidence intervals that cross OR threshold of 1.0 are not statistically significant at the alpha=0.05 level. All estimates presented are adjusted for other model variables. N=1,452. All responses collected between June-August 2020.

### Patient and Public Involvement

This study is a cross-sectional analysis of surveys conducted in 2020 using a multi-modal recruitment strategy. No patients were involved in our study. All analyses were conducted in collaboration with research partners in Dakar, Senegal so it was not appropriate to involve specific public communities or patients or in the design, conduct, reporting or dissemination plans.

## Results

A total of 1,452 Senegalese adults participated in our cross-sectional survey about COVID-19 (telephone=1,221; online=231). Table 1 presents demographics characteristics for the study sample. Table 2 presents univariate descriptive statistics for the three COVID-19 scales. Figure 1 presents a Forest plot of our logistic regression model predicting prevention behaviors.

**Table 2.**
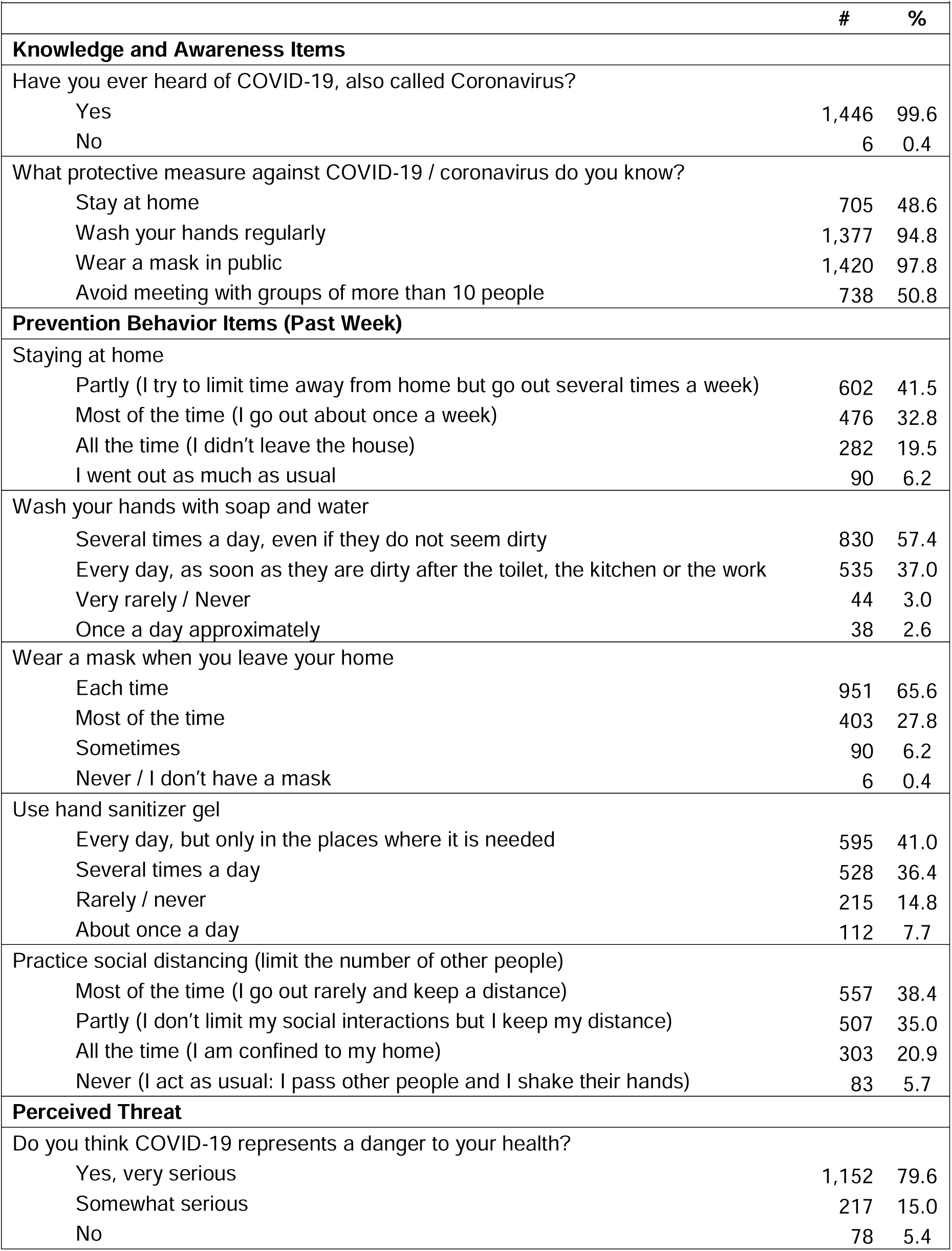
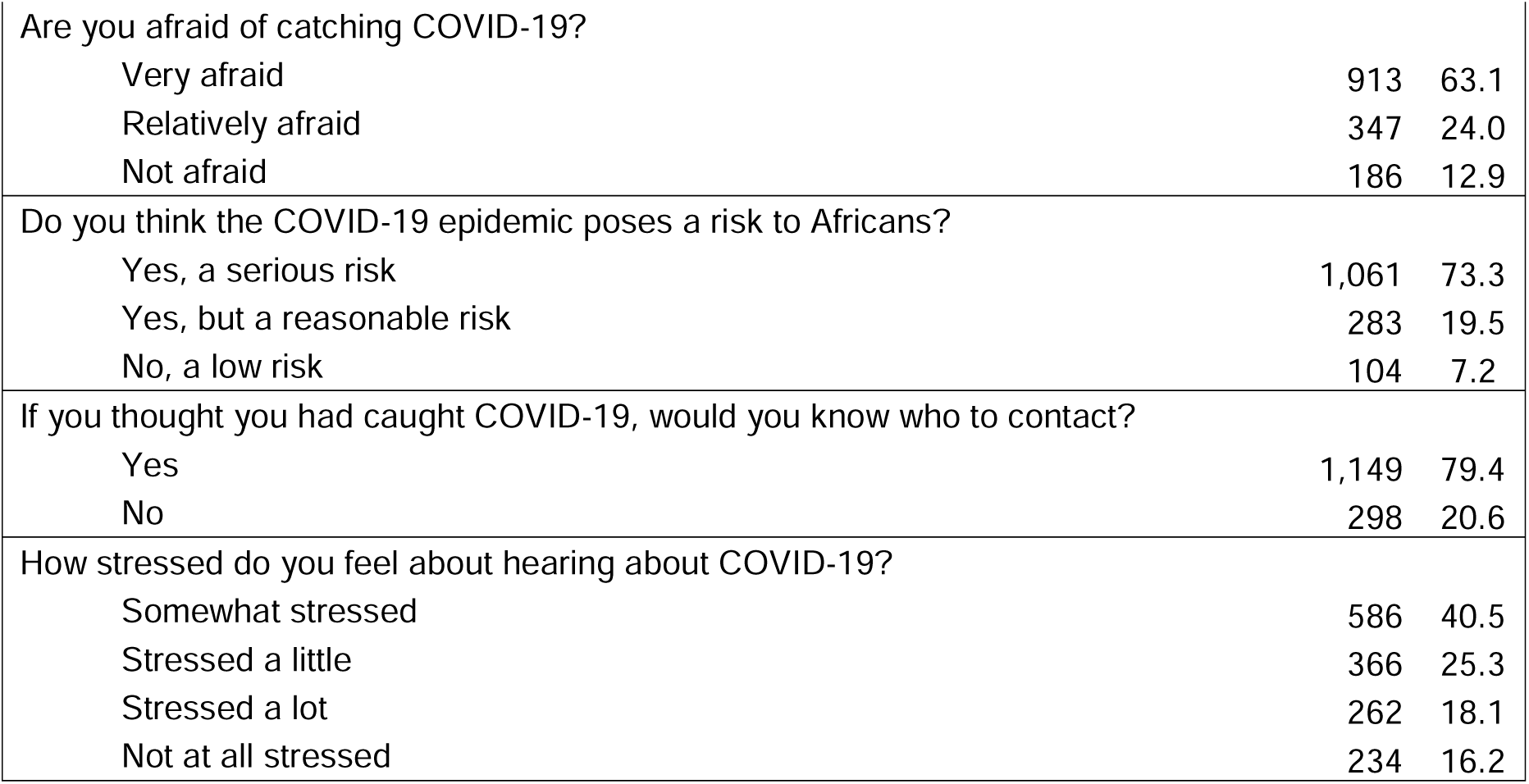
Frequency (#) and percent (%) for responses to COVID-19 questionnaire items. N=1,452 Senegalese adult participants. All responses collected between June-August 2020.

### Sample Characteristics

Most participants identified as Muslim (94.5%) and primarily lived in in an urban area (77.1%). Nearly half completed secondary education (high school or more; 49.9%). Most participants were single (55.4%). Participants’ ages ranges from 18-44 years, with a median age of 26 years and mean of 28.0 years (SD=7.6; mean and median data not presented). The sample was evenly split between females and males (50.1% versus 49.9%). A third of participants worked in the informal sector (e.g., craftsperson, restauranteur; 34.9%); another third were currently students (31.8%). Most participants completed the survey via phone rather than online, respectively 84.1% and 15.9%.

We identified some differences between our two recruitment methods – i.e., online versus telephone (data not presented). Compared to participants recruited online, participants recruited telephonically were more likely to be older on average (telephone=28.5 years versus online=25.0; p<.001), live in rural versus urban settings (telephone=23.0% rural versus online=4.4%; p<.001), and have less than college education (telephone=16.1% college-educated versus online=79.7%; p<.001). Half of telephone participants were male, compared to less than four in ten online participants (49.9% telephone versus 39.4% online; p=.003). Significant differences were also identified between recruitment methods for relationship status (p<.001), profession (p<.001), and religion (p<.001). Compared to telephone participants, online participants also had significantly higher COVID-19 knowledge and awareness (p<.001) and were significantly more likely to engage in prevention behaviors (p<.001).

### RQ1: In Senegal, what is the level of knowledge and awareness about COVID-19?

As shown in Table 2, of the 1,452 participants in our study sample less than 1% had not heard about COVID-19 (0.4%). More than nine in ten participants knew that hand washing (94.8%) and mask wearing in public (97.8%) were effective methods to prevent COVID-19 transmission and/or infection. About half were aware that staying at home (48.6%) or avoiding groups of more than 10 people (50.8%) were COVID-19 prevention recommendations. Nearly four in five participants (79.4%) said that they would know who to contact (e.g., testing, healthcare) if they suspected themselves to have had COVID-19. The median knowledge score was 12.5 out of 15 (range=0 [lowest] to 15 [highest]; mean=11.8 / SD=2.6; data not presented).

### RQ2: To what extent is COVID-19 perceived as a health threat?

As shown in Table 2, nine in ten participants (94.6%) perceived COVID-19 to be a very or somewhat serious danger to their health. Almost a quarter (73.3%) said that COVID-19 posed a serious risk to Africans. Median perceived threat score was 11.6 out of 15 (mean=10.9 /SD=3.5; data not presented).

### RQ3: What is the relationship between COVID-19 beliefs and prevention behaviors?

As shown in Table 2, nine in ten participants reported washing their hands multiple times per day (94.4%). Nearly eight in ten used hand sanitizer multiple times per day (77.4%). Approximately seven in ten participants wore masks every time they left their home (65.6%). Six in ten reported practicing social distancing all or most of the time (59.3%). Half of participants reported staying home all or most of the time (52.3%). Less than one in ten people went out as usual (6.2%) or ignored social distancing (5.7%). Less than one in one hundred participants never wore a mask in public (0.4%). The median prevention behaviors score was 11 out of 15 (mean=10.5/ SD=2.3; data not presented).

#### Regression Analysis

As shown in Figure 1 and Table 3, COVID-19 behaviors were significantly associated with COVID-19 beliefs and participant demographic attributes. We also observed differences in behaviors based on our participant recruitment method. Compared to participants recruited telephonically, participants recruited online were significantly more likely to socially distance (OR=3.33; p=<.001), wear masks (OR=3.26; p=.007), and stay home (OR=1.63; p=.013).

**Table 3.** Logistic regression models predicting COVID-19 prevention behaviors (aggregate + individually). N=1,452. All estimates presented are adjusted for other model variables. All responses collected between June-August 2020.

| *Predictor* | **<u>All Behaviors</u>** |  |  |  | **<u>Using Hand Sanitizer</u>** |  |  |  | **<u>Social Distancing</u>** |  |  |  |
| --- | --- | --- | --- | --- | --- | --- | --- | --- | --- | --- | --- | --- |
|  | *OR* | *95% CI* |  | *p* | *OR* | *95% CI* |  | *p* | *OR* | *95% CI* |  | *p* |
| Online participant | 1.93 | 1.29 | 2.87 | 0.001 | 1.62 | 0.94 | 2.78 | 0.079 | 3.20 | 2.05 | 5.00 | <.001 |
| COVID Perceived threat | 1.69 | 1.33 | 2.13 | <.001 | 1.69 | 1.28 | 2.22 | <.001 | 2.03 | 1.61 | 2.57 | <.001 |
| Rural | 1.49 | 1.12 | 1.98 | 0.006 | 0.67 | 0.50 | 0.91 | 0.010 | 1.36 | 1.03 | 1.81 | 0.032 |
| High school or above | 1.33 | 1.01 | 1.76 | 0.044 | 1.35 | 0.98 | 1.84 | 0.062 | 1.28 | 0.98 | 1.67 | 0.070 |
| Formal worker | 1.28 | 0.86 | 1.90 | 0.230 | 2.18 | 1.32 | 3.60 | 0.002 | 1.42 | 0.95 | 2.13 | 0.090 |
| Informal worker | 1.07 | 0.76 | 1.52 | 0.691 | 1.33 | 0.90 | 1.97 | 0.149 | 0.98 | 0.70 | 1.38 | 0.927 |
| COVID KAP | 0.96 | 0.72 | 1.29 | 0.798 | 1.47 | 1.03 | 2.10 | 0.035 | 0.89 | 0.66 | 1.18 | 0.411 |
| Single | 0.75 | 0.56 | 1.02 | 0.070 | 1.20 | 0.86 | 1.69 | 0.281 | 0.79 | 0.59 | 1.06 | 0.116 |
| 26 years or older | 0.75 | 0.56 | 1.00 | 0.054 | 1.19 | 0.86 | 1.64 | 0.287 | 0.78 | 0.59 | 1.03 | 0.077 |
| Male | 0.59 | 0.46 | 0.75 | <.001 | 0.64 | 0.49 | 0.84 | 0.001 | 0.62 | 0.49 | 0.79 | <.001 |
| *Predictor* | **<u>Staying Home</u>** |  |  |  | **<u>Hand Washing</u>** |  |  |  | **<u>Mask Wearing</u>** |  |  |  |
|  | *OR* | *95% CI* |  | *p* | *OR* | *95% CI* |  | *p* | *OR* | *95% CI* |  | *p* |
| Online participant | 1.62 | 1.09 | 2.42 | 0.018 | 2.48 | 0.89 | 6.96 | 0.084 | 3.25 | 1.31 | 8.04 | 0.011 |
| COVID Perceived threat | 1.62 | 1.29 | 2.03 | <.001 | 3.09 | 1.67 | 5.71 | <.001 | 2.81 | 1.64 | 4.82 | <.001 |
| Rural | 1.80 | 1.35 | 2.38 | <.001 | 1.52 | 0.81 | 2.86 | 0.189 | 2.17 | 1.12 | 4.19 | 0.021 |
| High school or above | 1.67 | 1.28 | 2.17 | <.001 | 0.85 | 0.49 | 1.45 | 0.546 | 0.82 | 0.50 | 1.36 | 0.448 |
| Formal worker | 0.98 | 0.67 | 1.44 | 0.918 | 1.54 | 0.67 | 3.53 | 0.311 | 2.44 | 1.03 | 5.75 | 0.042 |
| Informal worker | 0.89 | 0.64 | 1.25 | 0.510 | 1.12 | 0.58 | 2.16 | 0.733 | 1.10 | 0.60 | 1.99 | 0.760 |
| COVID KAP | 0.64 | 0.48 | 0.85 | 0.002 | 1.78 | 0.88 | 3.63 | 0.110 | 1.14 | 0.64 | 2.04 | 0.654 |
| Single | 0.85 | 0.64 | 1.13 | 0.260 | 0.93 | 0.50 | 1.71 | 0.809 | 0.73 | 0.41 | 1.29 | 0.277 |
| 26 years or older | 0.76 | 0.58 | 0.99 | 0.045 | 1.40 | 0.80 | 2.46 | 0.243 | 1.05 | 0.63 | 1.77 | 0.847 |
| Male | 0.49 | 0.39 | 0.62 | <.001 | 0.39 | 0.23 | 0.66 | <.001 | 0.61 | 0.39 | 0.97 | 0.037 |

Participants who reported high degrees of perceived threat about COVID-19 were significantly more likely to practice prevention behaviors in both the aggregate and individual models (OR=1.69; p<.001). Knowledge and awareness related to COVID-19 poorly predicted practicing prevention behaviors, although high levels of knowledge and awareness were positively associated with participants’ use of hand sanitizer (OR=1.47; p=.035). At the same time, higher levels of knowledge and awareness about COVID-19 was negatively associated with staying home (OR=0.64; *p=*.002).

Compared to participants with less than a high school degree, participants with high school education or above were more likely to stay home (OR=1.67; p<.001), practice social distancing (OR=1.28; p=.070), and use hand sanitizer (OR=1.35; p=.062). Compared to urban participants, rural participants were more likely to socially distance (OR=1.36; p=.032), stay home (OR=1.80; p<.001), and wear a mask (OR=2.17; p=.021), but less likely to user hand sanitizer (OR=0.67; p=.010). Use of hand sanitizer was more likely among formal workers compared to students (OR=2.18; p=.002). Compared to participants less than 26 years old, older participants (over 26) were significantly less likely to stay home (OR=0.76; *p*=.045).

Gender-based differences were observed for COVID-19 prevention behaviors. Men had 41% lower odds of practicing *all five* COVID-19 prevention behaviors compared to women (OR=0.59; p<.001). Men also reported lower adherence to COVID-19 prevention measures for each of the five behaviors individually. Compared to women, men were 61% less likely to wash their hands on a daily basis (p<.001), 51% less likely to report staying home (p<.001), 39% less likely to wear masks daily (p<.037), 38% less likely to practice social distancing (p<.001), and 37% less likely to use hand sanitizer (p=.001).

## Discussion

In our cross-sectional sample of Senegalese adults, we observed that the majority of participants practiced COVID-19 prevention behaviors on a daily basis. Mask wearing, hand washing, and use of hand sanitizer were most frequently reported. Social distancing and staying at home were also reported albeit to a lower degree. We also identified a range of psychosocial and demographic predictors for COVID-19 prevention behaviors. For example, men and women differed in the degree to which they practiced COVID-19 prevention behaviors. Participants who perceived COVID-19 to be a threat were significantly more likely to practice every prevention behavior measured than those who did not perceive COVID-19 to be a threat. This is in line with existing health behavior research regarding perceived threat as a necessary precursor to behavior change.[17,18]

### Prevention Behaviors

Men were less likely than women to report practicing any of the prevention behaviors that we studied. Other studies have shown that women are more likely to engage in preventive behaviors,[23,24] including towards COVID-19.[25,26] Men may also be more likely to engage in risk taking behavior,[27] whereas women are more risk averse.[28] Practicing COVID-19 prevention behaviors was also positively associated with living in a rural vs. urban area. It is possible that people residing in rural areas have an easier time implementing preventive behaviors such as staying at home and social distancing than people in more densely populated urban areas. Participants with at least a high school education were also more likely to engage in preventive behaviors. Educational differences in adoption of preventive health behaviors has previously been reported.[23,29] Demographic variations in practicing COVID-19 prevention behaviors indicate priority populations for future health communication campaigns. For example, people in urban areas may require more prompting and resources to implement preventive behaviors in the future.

Our results suggest that online communication will be an important strategy to build vaccine confidence and to continue supporting prevention behaviors especially as vaccine availability increases throughout the region. Respondents from the online sample demonstrated greater knowledge and awareness of COVID-19 and engaged in prevention behaviors more often compared to respondents in the telephone sample. This provides support for both online and on-the-ground messaging and communication to strengthen pro-prevention beliefs, attitudes, and behaviors.

### Health Beliefs

Our study findings comport with a robust body of evidence about COVID-19 knowledge, awareness, attitudes, and practices. In Senegal, we observed that knowledge, perceived risk, and practicing prevention behaviors for COVID-19 were very high in general – albeit with some demographic variations. A 2020 international review by Puspitasari and colleagues reported generally high knowledge and awareness about COVID-19, pro-prevention attitudes, and high adherence to COVID-19 prevention behaviors like handwashing and social distancing.[30] Closer to the West African and Senegalese contexts, Aduh and colleagues (2020) reported that in sub-Saharan Africa moderate to high levels of COVID-19 knowledge and perceived risk.[15]

Contrary to prior international research, we did not find an association between knowledge and engaging in prevention behaviors. Rather, perceived COVID-19 risk was a stronger predictor of prevention behaviors, which is also supported by previous international research.[2,31] Low perceptions of risk may result in low compliance with virus restrictions and adherence to prevention behaviors,[32–34] and therefore facilitates COVID-19’s spread. This confirms another recent study which recruited using social media from the United States, China, Taiwan, and Mexico.[2] Hsing and colleagues found that perceived COVID-19 severity and susceptibility – constructs that comprise perceived risk/threat in the Health Belief Model – were both predictors of hand washing and social distancing.[2] In our study based in Senegal, perceived risk not only predicted hand washing and social distancing, it also predicted staying home, using hand sanitizer, and mask wearing. These findings suggest that to prevent COVID-19 spread in Senegal and other parts of francophone West Africa, we need to focus more on perceived risk than knowledge.

### A Focus on Senegal

Senegal’s communication regarding COVID-19 has widely been considered successful.[35,36] In general, Senegalese participants had high levels of COVID-19 knowledge and awareness, perceived COVID-19 to be a threat both personally and across Senegal, and in most cases adopted known prevention behaviors on a daily or regular basis. Looking forward, as vaccines become more widely available in Senegal and many West African nations, inequities in access will make sustaining preventive behaviors paramount to continued control of the virus.[37] Our findings suggest that other nations should look to Senegalese communications about COVID-19 to learn messages that may resonate more effectively with their populations to increase prevention behaviors.

As has been stated, most participants reported moderate to high adherence to COVID-19 prevention behaviors. To explain our findings, we must consider lessons learned from previous regional epidemics and how they relate to evolving social contexts (e.g., norms, culture, population attributes).[20] COVID-19 in West Africa comes several years after the Ebola outbreak in 2014-2015, which tested the limits of the health systems. For example, although Senegal experiences a nationwide shortage of doctors – just 7 per 100,000 persons in 2020 – the nation has been comparatively effective at controlling COVID-19 infections. The first case of COVID-19 in Senegal, a country of 17 million, was reported in early March 2020, prompting the Ministry of Health to develop protocols to monitor and address cases within Senegal and at its borders and airports. Cases in Senegal remained low throughout Spring and Summer 2020, but like other nations rose considerably in the first months of 2021.[38,39]

Senegal’s success at controlling the virus throughout 2020 has been attributed to many factors, including a citizenry who has witnessed the devastating impacts of the Ebola epidemic. More broadly across West Africa, citizens may be more likely to comply with public health restrictions because of their experience with Ebola,[40] and this extends to individual behavior changes as well. For example, one study reported that handwashing was shown to increase in Senegal as a result of Ebola education campaigns.[41] In our analysis, we found that daily hand washing was the prevention behavior most often reported by participants. Future health campaigns may look at Ebola prevention messages to identify behaviors and information that may be reinforced in the context of COVID-19.

### Limitations

The current study is not without limitations. First, our findings may not be generalizable beyond Senegal, or more specifically Senegalese adults. Second, our recruitment strategy increased the potential for selection bias because participants were recruited online and on-the-ground in Senegal; thus, all participants, by the nature of the recruitment methods, had access to the internet and/or a cell phone. Selection bias is common in online and social media recruitment because recruited participants tend to be more well-educated and younger than the general population.[2,42] To address potential confounding between recruitment methods, we controlled for recruitment method in our multivariate regression modelling. At the same time, we believe that our multi-modal recruitment strategy bolsters the representativeness of our sample and is a key strength of our study. Had we excluded online participants, then our sample’s demographic characteristics would likely have differed. Conversely, had telephone participants been excluded then our sample would have skewed older, female, and educated – social media sample biases that we addressed through dual data collection methods.

Third and finally, we did not identify significant relationships for many factors in our regression model because our study was not statistically powered to detect significant differences. For example, COVID-19 knowledge was high in our sample and an insignificant predictor of prevention behaviors. A study with a larger sample may have been able to identify relationships between knowledge and behaviors. However, our study adapted pre-existing research panels and developed novel data collection instruments to capture information about COVID-19, and this may serve as a model for the responsiveness of ongoing scholarship to future global events.

## Conclusion

In the context of COVID-19, researchers must learn to apply traditional research methodologies online, such as on social media, to shed light on the etiology of COVID-19 in vulnerable and marginalized populations (Ali et al., 2020; Bavel et al., 2020). Such work creates evidence that may not only be applied to other places in developing regions but also enhance representation in research, which is crucial to creating tailored and effective interventions (Redwood & Gill, 2013). More broadly, research that creates evidence and informs policy about health issues like COVID-19 in West Africa and other parts of the developing world promotes global health equity and justice (Braveman & Gruskin, 2003; Cacari-Stone et al., 2014; Östlin et al., 2011).

Our study contributes to a growing body of literature about COVID-19 in Africa,[43,44] and is the first study to look specifically at knowledge and prevention behaviors in a francophone West African nation. In Senegal, we observed that knowledge and perceived risk of COVID-19 were very high in general. But, risk was a stronger and more influential predictor of COVID-19 prevention behaviors. COVID-19 prevention behaviors also varied by gender, education, location, and country. These findings suggest that in West Africa, nations should seek to elevate risk perceptions about COVID-19 and develop messaging to better target certain groups. Elevating the perceived risk of a population is a difficult issue because public health programs do not want to raise unnecessary alarm. Thus, messages around risk of COVID-19 should be accompanied by empowering messages of how individuals can protect themselves, their families, and their communities. Health campaigns should look backwards to successes with Ebola for improving health messaging about COVID-19, and may also look to replicate the successes of regional examples like Senegal.

## Supporting information

STROBE Checklist

## Data Availability

No data are available.

## Data availability statement

No data are available.

## Ethics statements

### Patient consent for publication

Not required.

### Research ethics approval

All study procedures and protocols were reviewed by and approved or determined to be exempt by Drexel University’s institutional ethical review board (Protocol ID: 00015164).

## Acknowledgements

We are grateful for support from our French and Senegalese collaborators in Dakar, Senegal at The African Health and Education Network (NGO RAES) and at Université Cheikh-Anta-Diop. Data collection was completed while the first (MK) and last author (PM) were affiliated with the Drexel University Dornsife School of Public Health, and while the second author (MB) was a graduate student at UCLA Fielding School of Public Health. Finally, we are thankful to all participants in this study.

## Contributors

All authors contributed to the editing and reviewing of this paper. MK, MB, DG, and PM were involved in conceptualizing this article. MF and RN led development of the data collection tool. MK and RN designed analyses for this manuscript, and MK conducted the analyses. MK, MB, DG, and PM wrote the paper. All authors reviewed, edited, and revised the paper.

